# Observation-process features are associated with larger domain shift in sepsis mortality prediction: a cross-database evaluation using MIMIC-IV and eICU-CRD

**DOI:** 10.64898/2026.04.05.26350209

**Authors:** Ryohei Yamamoto, Fan Wu, Laurine Kristin Sprehe, Ayesha Abeer, Leo Anthony Celi, Takeshi Tohyama

## Abstract

Clinical prediction models for sepsis frequently degrade when applied outside the development setting. Electronic health record data encode not only patient physiology but also observation processes such as measurement timing and frequency, which may be predictive within a site but unstable across sites. The contribution of these observation-process features to cross-site performance degradation has not been quantified. In this retrospective cohort study, we developed models for in-hospital mortality in adult intensive care unit (ICU) patients meeting Sepsis-3 criteria using Medical Information Mart for Intensive Care IV (MIMIC-IV) (n = 30,218; 16.3% mortality) and externally validated them in eICU Collaborative Research Database (eICU-CRD) (n = 31,403; 13.9% mortality). We compared seven prespecified model specifications representing physiologic summary strategies (a single aggregate severity score, most recent values, extreme values, and within-window variability), each evaluated with and without measurement counts as observation-process features. Models were fit using logistic regression and gradient-boosted trees. Internally, discrimination improved with more detailed physiologic summaries and measurement counts (logistic regression area under the receiver operating characteristic curve [AUROC] from 0.819 to 0.834). In external validation, performance drops were larger for specifications using more complex physiologic representations. Adding measurement counts was associated with larger domain shift (AUROC change, −0.047 versus −0.082 with counts in logistic regression). External calibration deteriorated progressively, with calibration slopes decreasing from 1.007 for the simplest model to 0.417 for the most complex specification in logistic regression. Gradient-boosted trees showed smaller incremental degradation from measurement counts but still exhibited domain shift in complex specifications. Inclusion of observation-process features in sepsis mortality prediction models was associated with improved internal discrimination but worse external calibration and transportability. These findings highlight that feature engineering decisions involve a tradeoff between internal performance and external generalizability, and that calibration assessment provides the most sensitive indicator of reduced transportability.

**Author Summary:** We asked whether the way physiologic data are summarized and how clinical measurements are recorded in electronic health records can affect how well prediction models perform at new hospitals. Using data from over 60,000 intensive care patients with sepsis across two large databases, we built models to predict in-hospital death using distinct physiologic summary strategies of varying complexity. First, we found that extending physiologic summaries from simple aggregate scores to include extreme values and variability ranges was associated with improved predictions within the hospital where the model was developed, but with poorer calibration and reduced transferability when the model was applied to other hospitals. Second, we found that further incorporating observation-process features, such as measurement frequency, was also associated with improved internal discrimination but with worse external calibration and transferability. These patterns likely arise because richer summaries and measurement patterns capture not only patient physiology but also facility-specific clinical workflows and care processes. Our results suggest that model developers should carefully consider whether the data features they use capture stable biological signals or hospital-specific practices, and that checking whether predicted risks remain well-calibrated across settings is the most important step before deploying a model in a new hospital.

## Introduction

Clinical prediction models often degrade when applied outside the development setting because the data generating process differs across populations and sites, a phenomenon commonly referred to as dataset shift or domain shift [1,2]. Such shifts can arise from differences in case mix, baseline risk, measurement practices, and care delivery, and they may compromise both discrimination and calibration. External validation is therefore essential when models are intended for use beyond the original development environment [3,4].

This challenge is particularly consequential in critical care, where prediction models are used for high-stakes decisions. In sepsis, external validation has repeatedly shown that apparently strong internal performance does not guarantee reliable risk estimation in routine care settings [5]. A prominent example is the independent external validation of a widely implemented commercial sepsis prediction model, which demonstrated poor discrimination and poor calibration [6]. Systematic reviews similarly highlight frequent limitations in validation practices and generalizability for sepsis prediction models [7,8]. A plausible, underappreciated mechanism is that electronic health record data encode not only physiology but also the observation process. Measurement timing and frequency are shaped by clinician judgment and local workflows, creating informative observation patterns that may be predictive within a site but unstable across sites [9–12]. Related work also shows that visit and measurement processes can induce bias and limit the portability of inferences drawn from routine health data [13,14]. Consistent with this concern, recent intensive care unit (ICU) prediction studies have begun to include observation-process features such as measurement frequency, missingness rates, or measurement counts as model inputs, which may improve within-site discrimination but can also make prediction and transportability more sensitive to cross-site differences in care and documentation practices [15,16].

These considerations motivate a deployment-relevant hypothesis: enriching physiologic summaries in ways that increasingly capture observation-process characteristics may be associated with improved internal discrimination alongside worse external calibration and reduced transportability. In this study, we quantify these tradeoffs by comparing prespecified feature sets representing distinct summary strategies under external validation and by evaluating both discrimination and calibration.

## Materials and methods

### Study design and data sources

This retrospective cohort study developed prediction models in Medical Information Mart for Intensive Care IV (MIMIC-IV) and externally validated them in eICU Collaborative Research Database (eICU-CRD) [17–20]. Both are publicly available, de-identified ICU databases derived from electronic health records and include patient demographics, diagnoses, physiologic measurements, laboratory tests, treatments, and outcomes. We used the two databases to evaluate model transportability under cross-dataset differences in patient mix, measurement practices, and documentation.

#### Derivation cohort (MIMIC-IV)

MIMIC-IV version 3.1 contains de-identified clinical data from ICU admissions at a single academic tertiary care center (Beth Israel Deaconess Medical Center) in Boston, Massachusetts, United States between 2008 and 2022 [17,18]. The derivation cohort was drawn from adult medical, surgical, cardiac, and neurological ICUs at this center. The database includes time-stamped bedside vital signs and laboratory measurements recorded during ICU care, enabling construction of features that incorporate both physiologic values and measurement intensity over time.

#### External validation cohort (eICU-CRD)

The eICU-CRD contains de-identified clinical data from adult ICU admissions across 208 hospitals in the United States during 2014 to 2015 [19,20]. These hospitals represent a geographically diverse range of community and academic medical centers, providing a multi-center external validation cohort distinct from the derivation setting. Because data recording practices, coding, and completeness can vary across hospitals, eICU-CRD provides a pragmatic test of external validity, including calibration performance under differences in measurement and documentation.

### Study population

We included adult patients (aged 18 years or older) who met Sepsis-3 criteria at ICU admission. Sepsis-3 was operationalized as suspected infection with an acute increase in the Sequential Organ Failure Assessment (SOFA) score of 2 points or more [21–23]. Suspected infection was identified by the co-occurrence of antibiotic administration. We required an ICU length of stay of at least 24 hours to ensure availability of time-updated clinical measurements. To ensure one observation per patient, only the first ICU admission meeting sepsis criteria was included. ICU stays involving patients younger than 18 years or lasting less than 24 hours were excluded.

### Outcome

The primary outcome was in-hospital mortality, defined as death during the same hospital admission as the index ICU stay, without a prespecified time horizon. Mortality status was obtained from the administrative discharge disposition field. Because the outcome was derived from the same administrative field for all patients, outcome assessment did not require subjective interpretation and was applied consistently across sociodemographic groups. Outcome assessment was not blinded to predictor values because the study used routinely collected retrospective data.

### Predictor selection rationale

To facilitate transparent comparisons across model specifications, we restricted candidate predictors to variables included in the Acute Physiology and Chronic Health Evaluation III (APACHE III) framework and available in both databases [24]. Specifically, we used APACHE III related demographics, comorbidity categories, and physiologic measurements collected during routine ICU care. All predictors in the harmonized dataset were prespecified and considered. No data-driven pre-selection such as univariable screening was performed before model fitting.

### Definitions and feature construction

All predictors were derived from routinely collected data within the first 24 hours after ICU admission, and models were designed to use only information available up to 24 hours to predict subsequent in-hospital mortality. Candidate variables included age (years), an APACHE III style comorbidity category, and 18 continuous physiologic variables measured during routine ICU care: vital signs (heart rate, mean arterial pressure, temperature, respiratory rate, and Glasgow Coma Scale), laboratory values (white blood cell count, hematocrit, sodium, glucose, blood urea nitrogen, creatinine, bilirubin, and albumin), blood gas parameters (pH, partial pressure of arterial oxygen [PaO2], partial pressure of arterial carbon dioxide [PaCO2], fraction of inspired oxygen [FiO2], and alveolar-arterial gradient), and urine output. Glasgow Coma Scale was assessed at each time point by summing eye, verbal, and motor component scores, and then summarized within the 24-hour window.

For each of the 18 continuous variables, we computed summary features within the 24-hour window as follows. We derived the latest value (most recent measurement) for all 18 variables. We also derived minimum and maximum values for all 18 variables. In addition, we calculated the measurement count (number of recorded measurements) for each of the 18 variables, which served as a proxy for measurement intensity and was treated as an observation-process feature that may reflect differences in observation and documentation practices across settings in addition to patient state.

For model specifications that included variability features, we computed within-window variability as the difference between maximum and minimum values (max minus min) for each of the 18 physiologic variables.

We also calculated the APACHE III score using the worst physiologic values within the first 24 hours, following the original methodology.

Treatments and care status during the first 24 hours of the ICU stay were recorded as descriptive variables, including mechanical ventilation status and acute renal failure, but were not included as predictors in the models. Models were designed to predict mortality regardless of treatment decisions, and the study population was not restricted by treatments received. Predictor assessment was not blinded given the retrospective design using routinely collected data.

### Statistical analysis

We developed prespecified model specifications to quantify how inclusion of measurement intensity as an observation-process proxy is associated with internal performance and external transportability under cross-dataset domain shift. Model development was performed in MIMIC-IV and external validation was performed in eICU-CRD. The external validation cohort was not used for model training, imputation fitting, or hyperparameter tuning.

#### Data partitioning and analysis sets

The derivation cohort was split into training (60%) and internal validation (40%) sets using stratified random sampling to preserve outcome proportions. A 60/40 split was chosen to provide a larger internal validation set, enabling more precise estimation of calibration metrics, which were of primary interest for assessing transportability. The entire eICU-CRD cohort served as the external validation set.

#### Model specifications

To assess how observation-process features are associated with internal performance and external transportability under domain shift, we prespecified seven model specifications. To facilitate direct comparisons, candidate predictors were restricted to variables aligned with the APACHE III framework and available in both databases. Specifications differed by how physiologic information from the first 24 hours after intensive care unit admission was summarized and by whether measurement counts were included as an observation-process feature. The physiologic summary strategies (most recent values, minimum/maximum, and within-window variability) are not strictly nested; they represent alternative representations of the same underlying physiologic time series. Within each strategy, paired specifications with and without measurement counts allow direct evaluation of the incremental contribution of observation-process features. A complete variable list for each specification is provided in S1 Table.

#### Model types and preprocessing

For each feature set, we fit logistic regression with default L2 regularization as the primary model to improve coefficient stability across specifications and to provide a transparent baseline for assessing external transportability under domain shift. Preprocessing, including imputation, scaling, and one-hot encoding, was implemented within scikit-learn pipelines and fit on the derivation training data only to prevent data leakage. For gradient-boosted trees (XGBoost) [25], hyperparameters were tuned using stratified five-fold cross-validation and randomized search within the derivation training data.

Treatments received during the first 24 hours of the ICU stay were recorded but were not included as predictors, and the cohort was not restricted by treatment received. Predictor assessment was not blinded given the retrospective design.

#### Model evaluation

We evaluated discrimination using area under the receiver operating characteristic curve (AUROC) and area under the precision-recall curve (AUPRC). Calibration was assessed using calibration curves, Brier score, and calibration intercept and slope. Apparent overfitting was summarized as the difference between training and internal validation AUROC. We computed 95% confidence intervals (CI) for AUROC and AUPRC using nonparametric percentile bootstrapping with 1,000 resamples; bootstrap samples containing only one outcome class were excluded.

For external validation, predicted probabilities were generated by applying the trained pipelines from the derivation data directly to eICU-CRD without adaptation. We did not perform model updating or recalibration using the external data, in order to quantify raw transportability across model specifications.

#### Heterogeneity, class imbalance, missing data, and sample size

Because the derivation cohort was single-center, we did not estimate heterogeneity of model parameters across clusters in the development setting. For descriptive fairness reporting, we assessed subgroup performance stratified by race using three models: Model 1 (APACHE III only), the best-performing specification without measurement counts, and its count-augmented counterpart. Performance was evaluated in both the internal validation and external validation cohorts. Given moderate class imbalance, we did not apply resampling or class-weighting methods.

Missingness was examined for each variable. For logistic regression, numerical features were imputed using multivariate imputation by chained equations (MICE) with Bayesian ridge regression as the estimator within the pipeline, whereas for XGBoost we used median imputation. Measurement count features were set to zero when no measurements were recorded within the first 24 hours; this approach does not distinguish between variables for which measurement was not clinically indicated and variables that were indicated but not recorded, which may introduce information bias if the reasons for non-measurement differ across settings. Missing data handling was applied uniformly across all patients.

Sample size was determined by the number of eligible patients available in each database. No formal sample size calculation was performed.

## Declarations

### Ethics approval and consent to participate

All records in MIMIC-IV and eICU-CRD are de-identified in accordance with the Health Insurance Portability and Accountability Act Safe Harbor provisions. Because this study used only publicly available de-identified data, institutional review board approval and informed consent were not required. Access to both databases requires completion of the relevant data use agreements and human subjects research training.

### Competing interests

The authors have declared that no competing interests exist.

### Protocol and registration

A formal protocol was not prepared, and the study was not registered in a prediction model registry.

### Data availability

Study data are available via PhysioNet: MIMIC-IV (https://physionet.org/content/mimiciv/) and eICU-CRD (https://physionet.org/content/eicu-crd/). Access requires credentialing and acceptance of the data use agreements.

### Code availability

Code used to generate the study cohorts, analyses, tables, and figures is publicly available at https://github.com/ryohei-hey/sepsis-observation-process-domain-shift.

### Patient and public involvement

Patients and members of the public were not involved in the design, conduct, or dissemination of this study.

## Results

### Participant flow

We identified 43,705 ICU stays meeting Sepsis-3 criteria in MIMIC-IV and 43,572 in eICU-CRD (Fig 1). After restricting to each patient’s first sepsis ICU admission and excluding ICU stays shorter than 24 hours, the final cohorts included 30,218 patients in MIMIC-IV (derivation) and 31,403 patients in eICU-CRD (external validation).

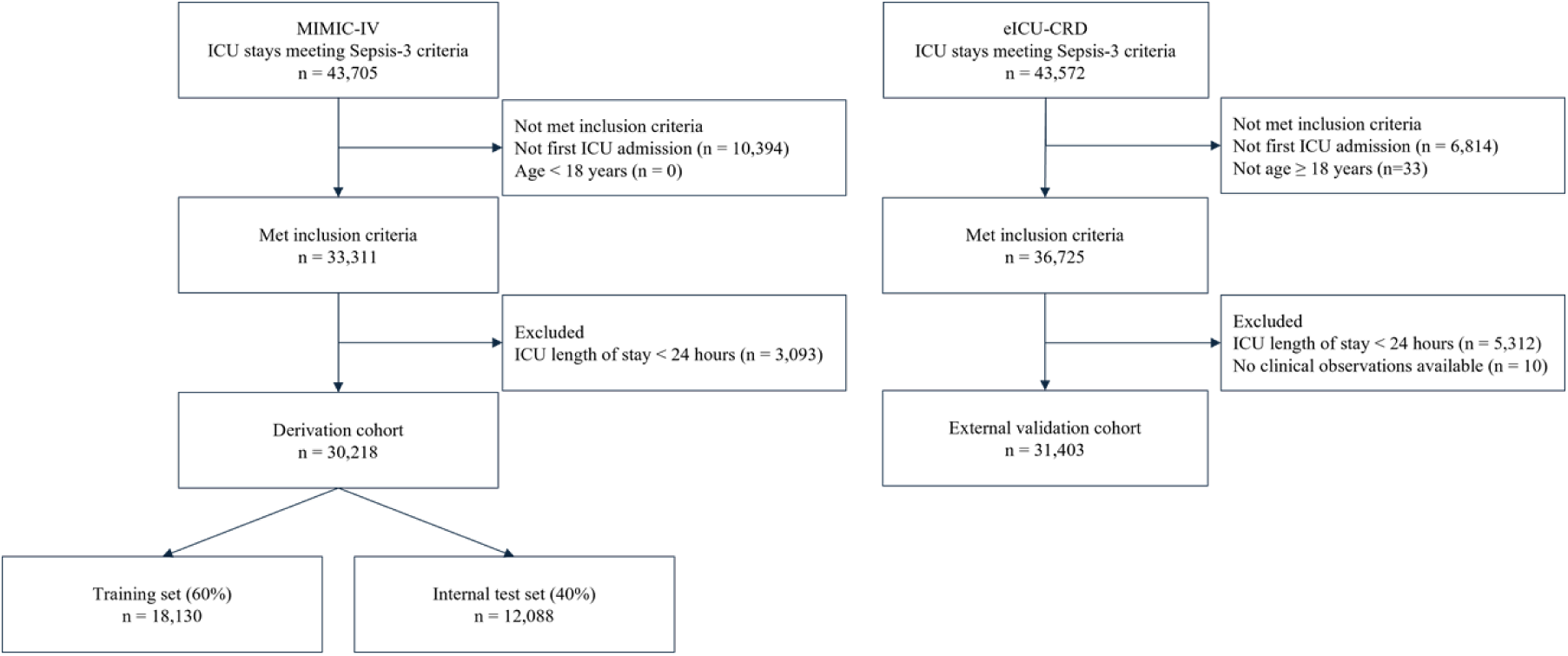

### Baseline characteristics

Baseline characteristics are shown in Table 2. Patients were of similar age across cohorts (mean 66.0 years in MIMIC-IV and 65.2 years in eICU-CRD), and sex distributions were comparable. Illness severity was higher in the derivation cohort, with a higher median APACHE III score (70 [interquartile range (IQR) 54 to 90] vs 65 [49 to 85]). The derivation cohort included 30,218 patients and the external validation cohort included 31,403 patients; in-hospital mortality was 16.3% (4,926 deaths) in MIMIC-IV and 13.9% (4,371 deaths) in eICU-CRD.

**Table 1.** Summary of model specifications used for the primary comparisons. **Legend:** Models 3, 5, and 7 included measurement counts as the primary observation-process feature, operationalized as the number of recorded measurements for each physiologic variable during the first 24 hours. The three physiologic summary strategies (most recent values, minimum/maximum, and within-window variability) are not strictly nested but represent alternative representations; the paired design (with and without measurement counts) isolates the incremental association of observation-process features with changes in internal discrimination and external calibration within each strategy.

| Model | Physiologic information | Measurement counts | Other predictors |
| --- | --- | --- | --- |
| 1 | APACHE III score only | No | None |
| 2 | Most recent values | No | Age, urine output, comorbidity category |
| 3 | Most recent values | Yes | Age, urine output, comorbidity category |
| 4 | Minimum and maximum values | No | Age, urine output, comorbidity category |
| 5 | Minimum and maximum values | Yes | Age, urine output, comorbidity category |
| 6 | Within-window variability (max minus min) | No | Age, urine output, comorbidity category |
| 7 | Within-window variability (max minus min) | Yes | Age, urine output, comorbidity category |

**Table 2.**
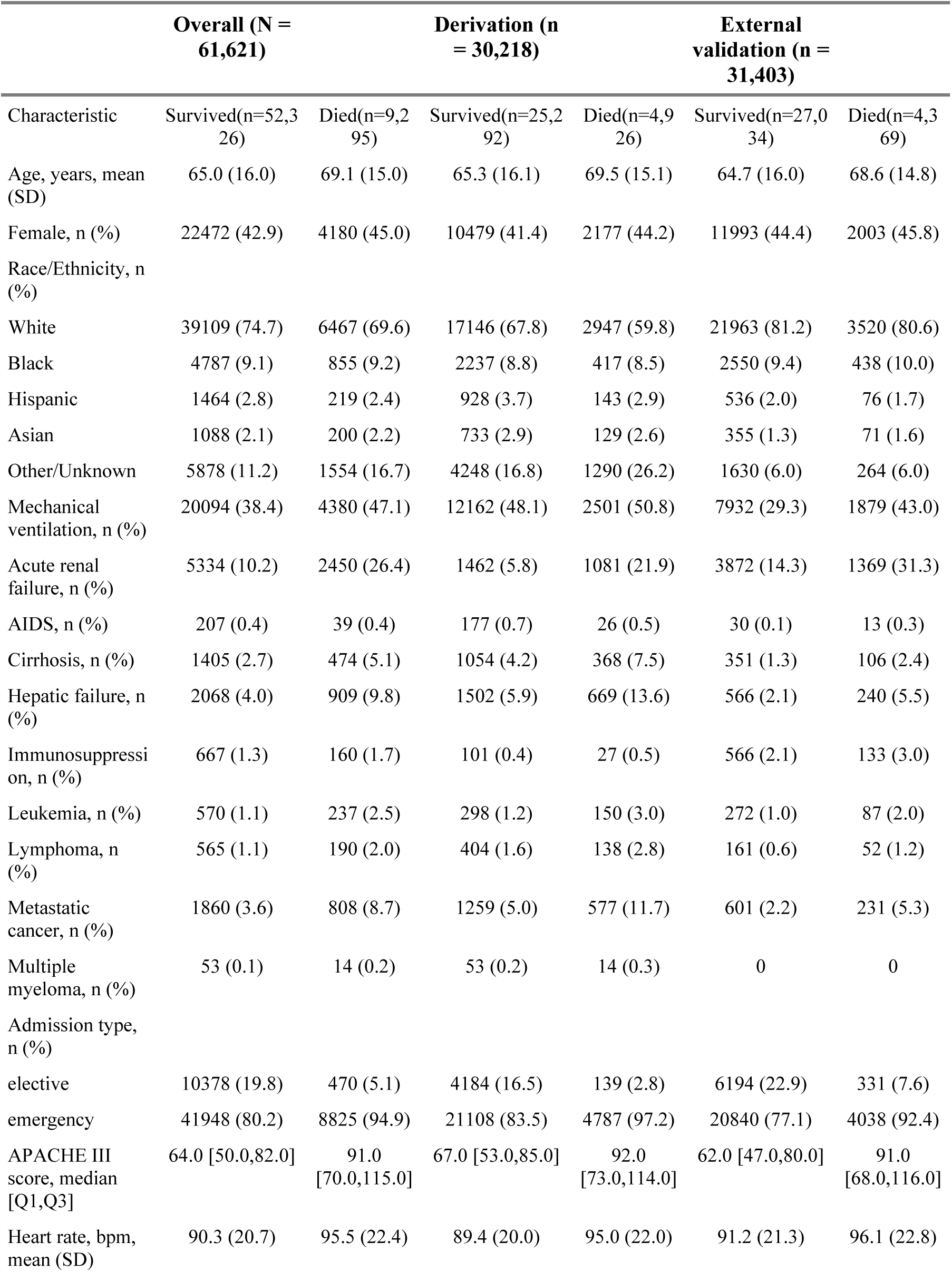
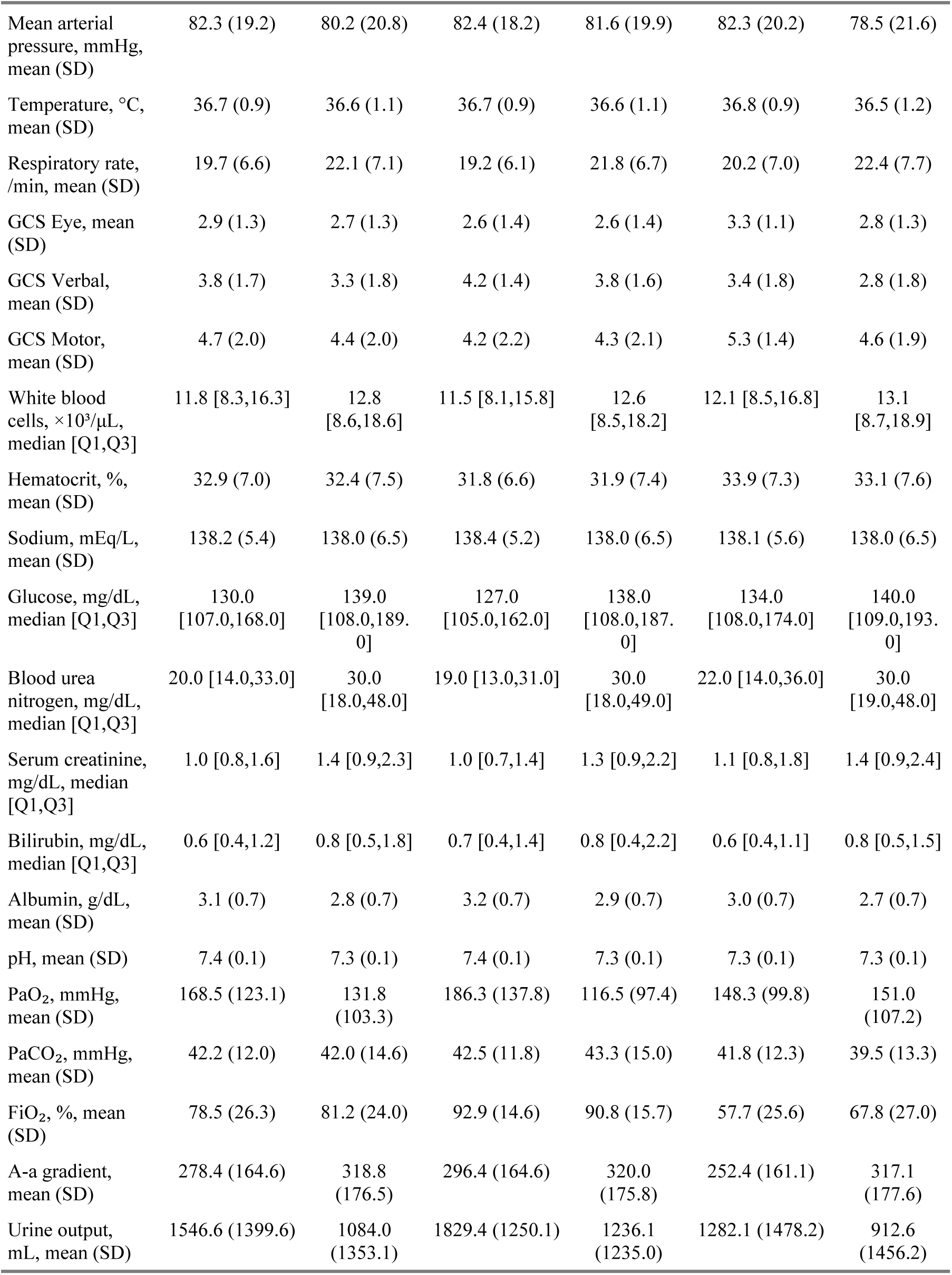
Baseline characteristics of patients with sepsis stratified by cohort and hospital mortality.

Several predictors differed between cohorts. Compared with eICU-CRD, MIMIC-IV had higher rates of mechanical ventilation (48.5% vs 31.2%) and hepatic-related comorbidity (hepatic failure 7.2% vs 2.6%; cirrhosis 4.7% vs 1.5%), as well as higher prevalence of metastatic cancer (6.1% vs 2.6%), whereas acute renal failure was less frequent (8.4% vs 16.7%). Mean fraction of inspired oxygen was higher in MIMIC-IV (92.5% vs 59.4%), consistent with differences in data recording conventions between databases.

Distributions of individual predictors are shown in S1 Fig, and measurement count distributions are shown in S2 Fig. Measurement counts differed markedly between cohorts, with more frequent measurements recorded in MIMIC-IV across most variables.

### Internal and external validation performance

The derivation cohort was split into a training set (n = 18,130) and an internal validation set (n = 12,088) for model fitting and internal validation. We evaluated seven prespecified model specifications (Models 1 to 7) representing distinct physiologic summary strategies of varying complexity (S1 Table). Table 3 presents the performance comparison between the internal validation set (held-out MIMIC-IV, 40%) and the external validation cohort (eICU-CRD) for logistic regression and XGBoost across all specifications.

**Table 3.**
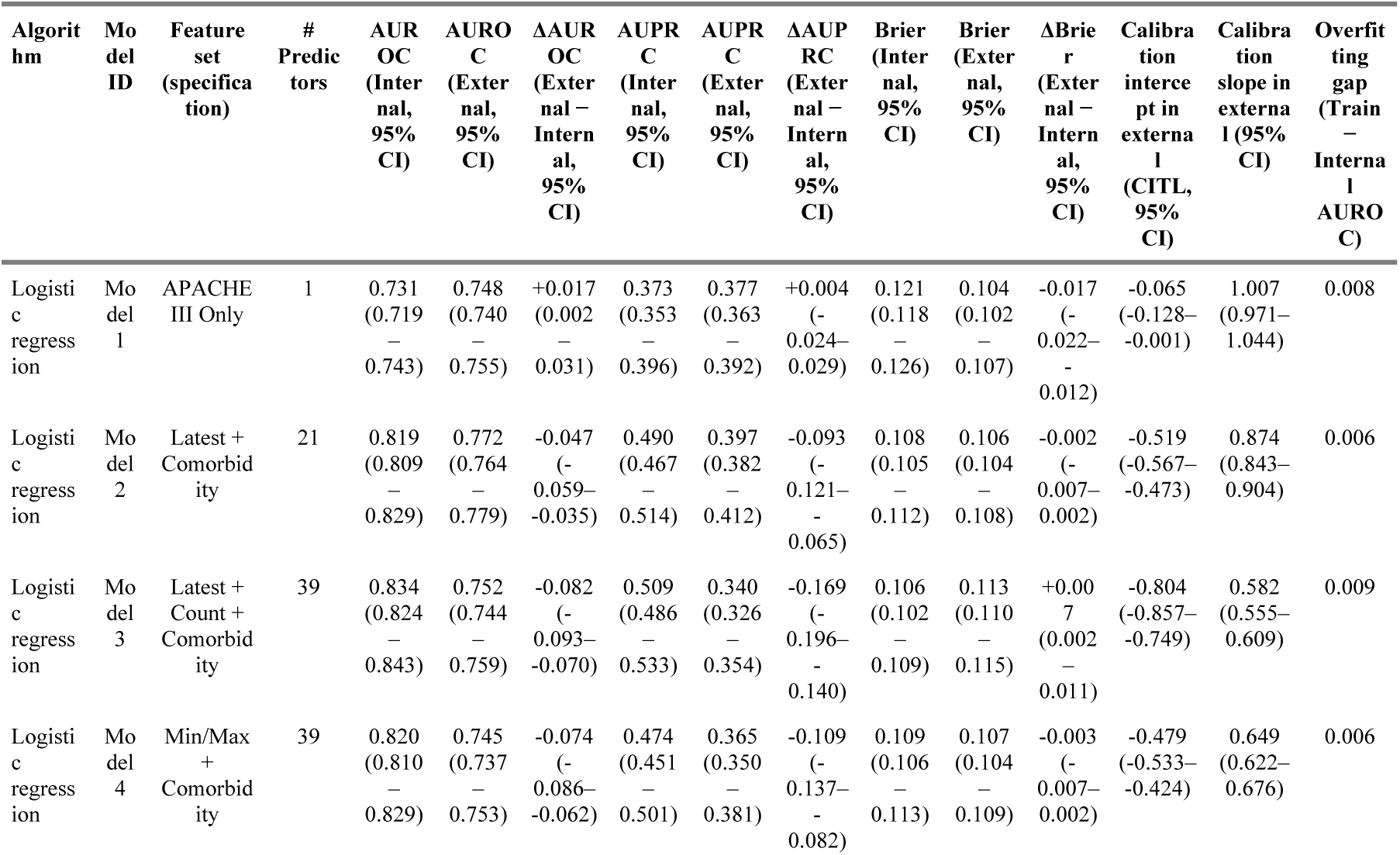
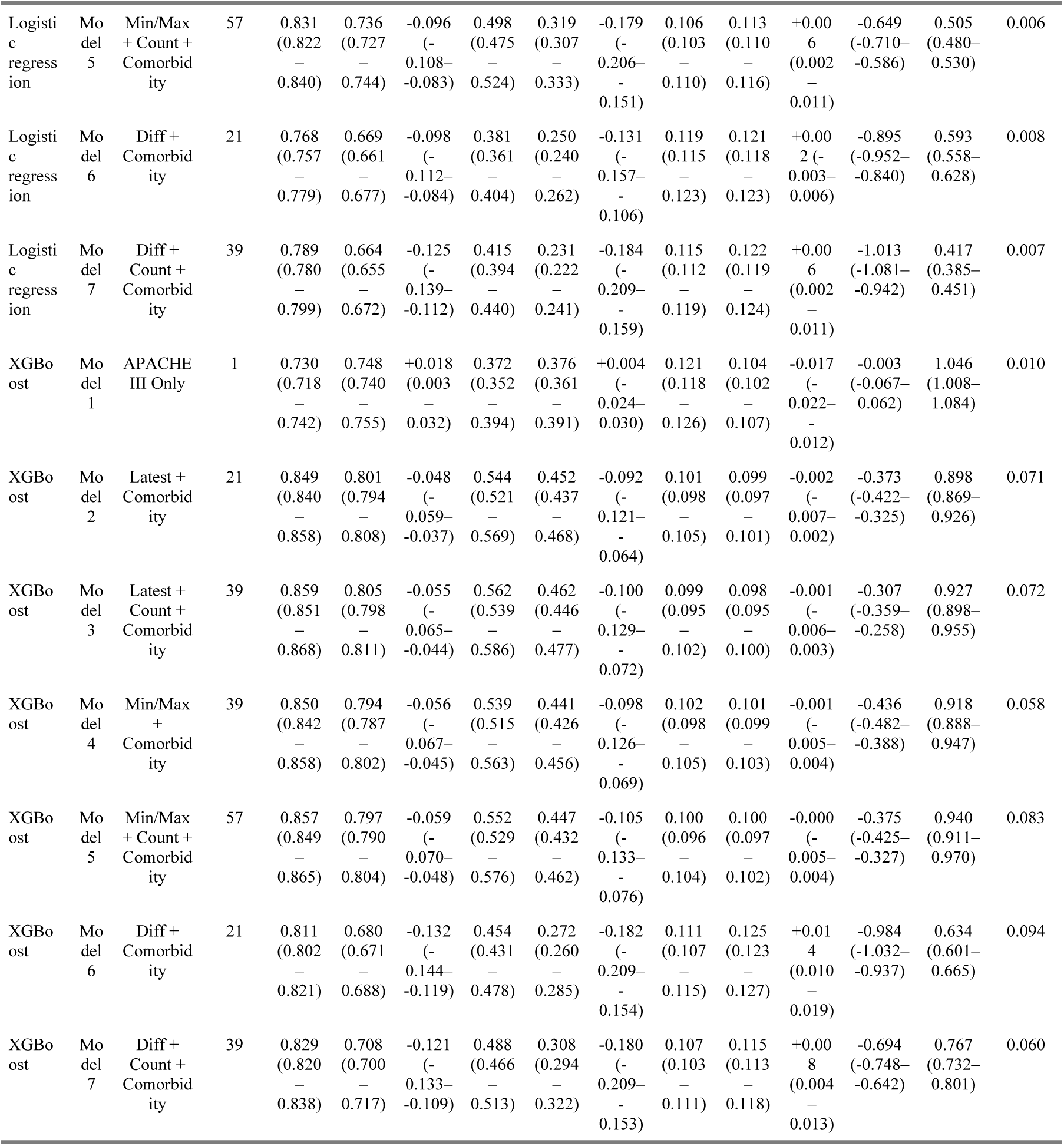
Internal-to-external validation performance (domain shift) across model specifications and algorithms.

#### Discrimination

In internal validation, the APACHE III baseline (Model 1) had AUROC around 0.73 in both algorithms. Internal discrimination was higher for specifications using more detailed physiologic summaries, and measurement count features were associated with further increases in AUROC, for example in logistic regression from 0.819 (Model 2) to 0.834 (Model 3). In external validation, logistic regression AUROC for Model 2 was 0.772 and for Model 3 was 0.752. In XGBoost, the highest external AUROC was observed for Model 3 (0.805). Full discrimination results are reported in Table 3, with ROC and precision-recall curves in S3–S6 Figs.

#### Domain shift

Fig 2 summarizes domain shift as ΔAUROC (external minus internal), where more negative values indicate a larger drop in discrimination in external validation. For logistic regression, ΔAUROC became more negative across specifications using more complex physiologic representations, from +0.017 in Model 1 to −0.125 in Model 7. Within each paired specification, models including measurement count features showed larger drops: Model 3 versus Model 2 (−0.082 vs −0.047), Model 5 versus Model 4 (−0.096 vs −0.074), and Model 7 versus Model 6 (−0.125 vs −0.098).

**Fig 2.**
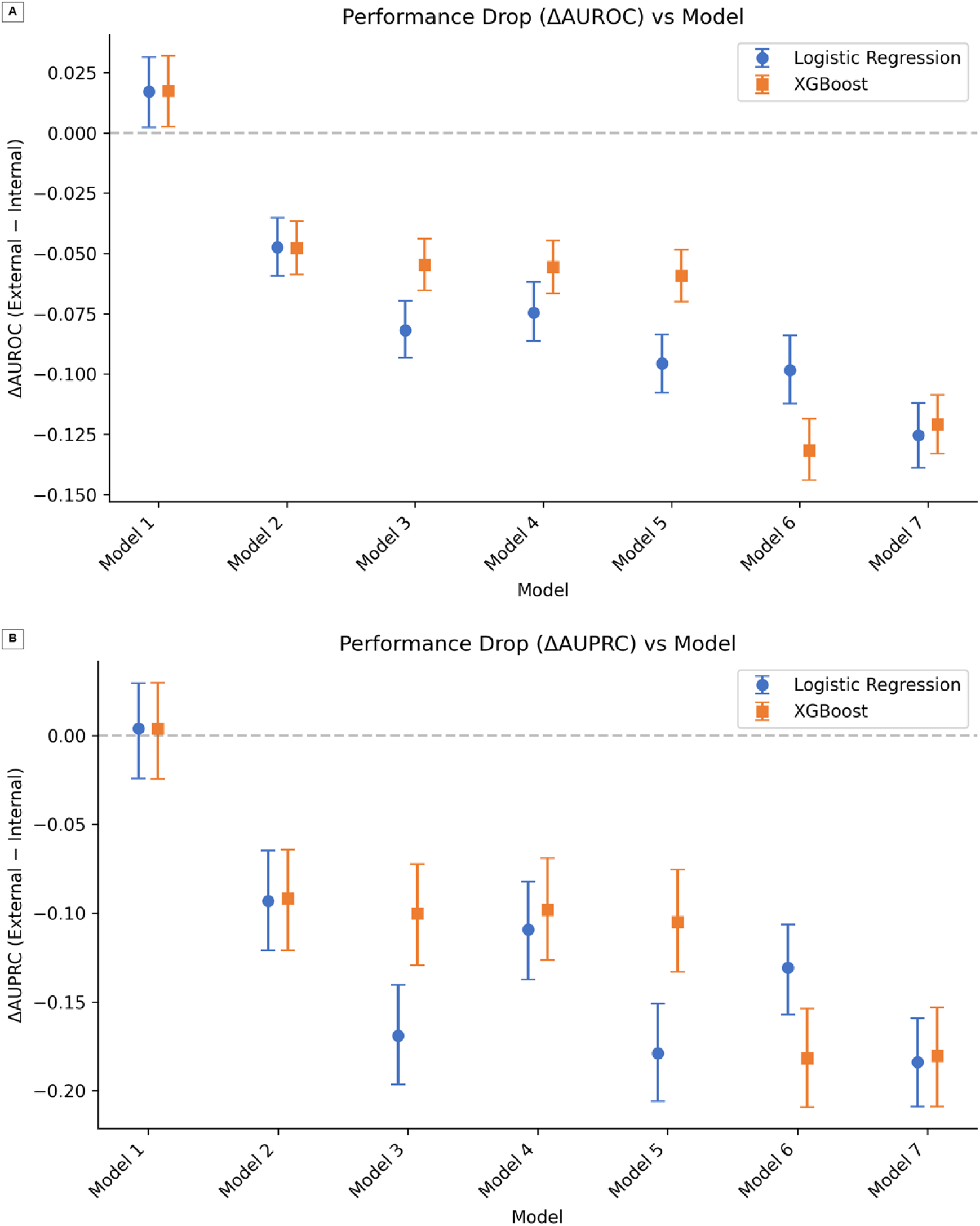
Performance drop from internal to external validation across model specifications Fig 2. Performance drop from internal validation to external validation across model specifications. (A) ΔAUROC and (B) ΔAUPRC, with 95% bootstrap CIs. Model 1 (APACHE III only) showed no degradation; models including measurement count features (Models 3, 5, 7) were consistently associated with larger domain shift relative to their counterparts without counts (Models 2, 4, 6).

For XGBoost, ΔAUROC also became more negative with increasing complexity, with the largest drop in Model 6 (−0.132). The effect of measurement count features on ΔAUROC was small for the latest-value and min/max specifications (Model 3 vs 2: −0.055 vs −0.048; Model 5 vs 4: −0.059 vs −0.056), whereas the variability specification showed a large drop regardless of whether counts were included (Model 6: −0.132; Model 7: −0.121). To help distinguish the role of feature type from dimensionality, we compared specifications with the same number of input features but different physiologic representations. Model 2 (latest values, 21 features) and Model 6 (within-window variability, 21 features) had the same dimensionality, yet Model 6 showed a substantially larger ΔAUROC in logistic regression (−0.098 vs −0.047). Similarly, Model 3 (latest values plus counts, 39 features) and Model 4 (min/max values, 39 features) had the same dimensionality; Model 3, which included measurement counts, showed a larger ΔAUROC (−0.082 vs −0.074). These dimensionality-matched comparisons suggest that the type of feature, particularly whether it encodes observation-process or within-window summary information, is associated with domain shift beyond the effect of increasing the number of model inputs.

Differences in AUROC and AUPRC with 95% bootstrap CIs are shown in Fig 2.

#### Calibration

External calibration varied across model specifications and algorithms (Figs 3–4; Table 3). In logistic regression, external calibration was less favorable for specifications using more complex physiologic representations. Calibration slope decreased from 1.007 in Model 1 to 0.417 in Model 7, and the calibration intercept became more negative from −0.065 to −1.013. Within paired specifications, models including measurement count features had lower calibration slopes and more negative calibration intercepts on external validation than their counterparts without counts, for example Model 2 versus Model 3 (slope 0.874 vs 0.582; intercept −0.519 vs −0.804) and Model 4 versus Model 5 (slope 0.649 vs 0.505; intercept −0.479 vs −0.649).

**Fig 3.**
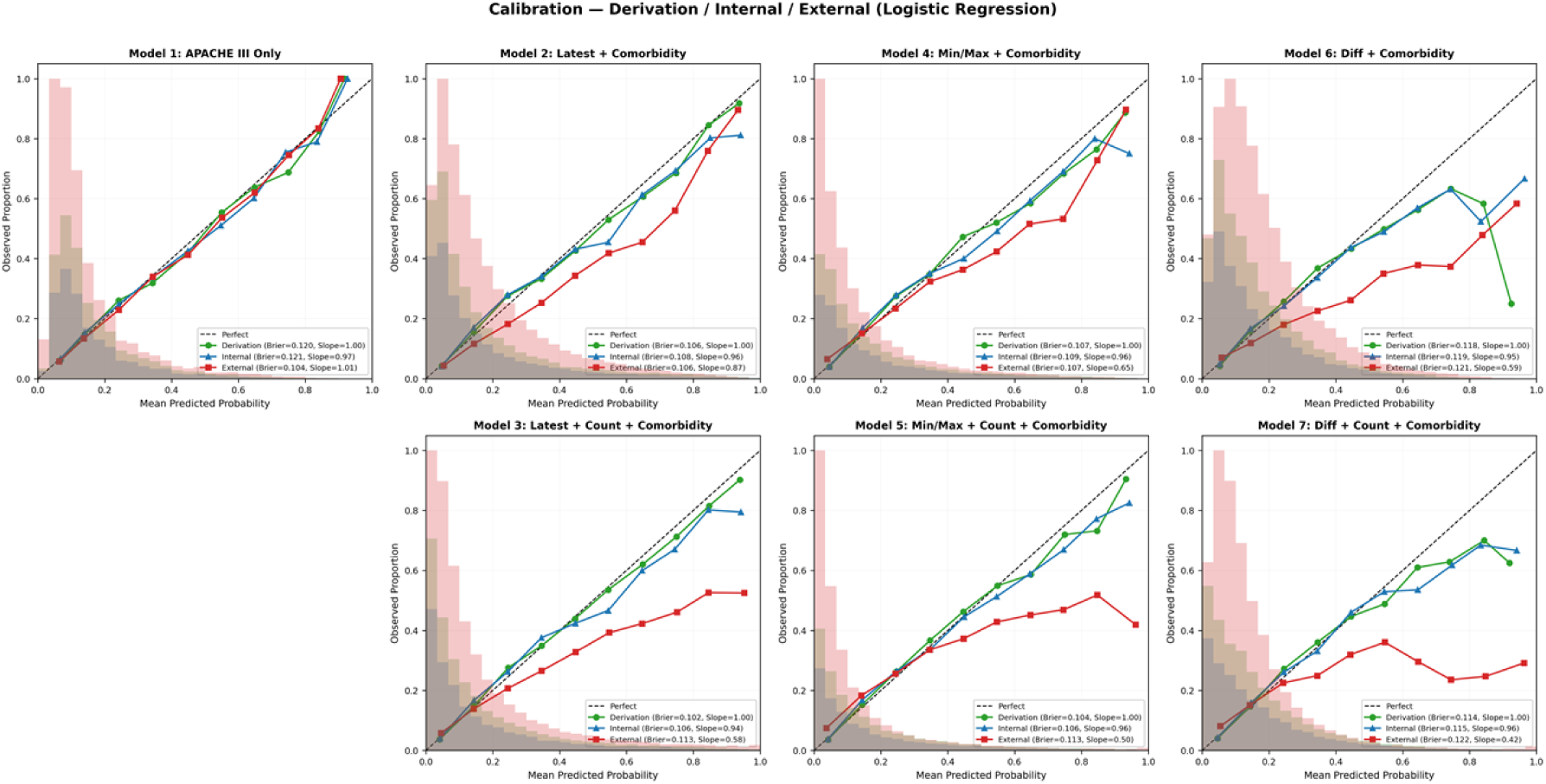
Calibration plots for logistic regression models on derivation, internal validation, and external validation sets Calibration plots for logistic regression Models 1–7 on derivation (training), internal validation, and external validation sets. Upper row: models without count features; lower row: paired models with count features. Models incorporating count features showed progressively worse calibration on external validation.

**Fig 4.**
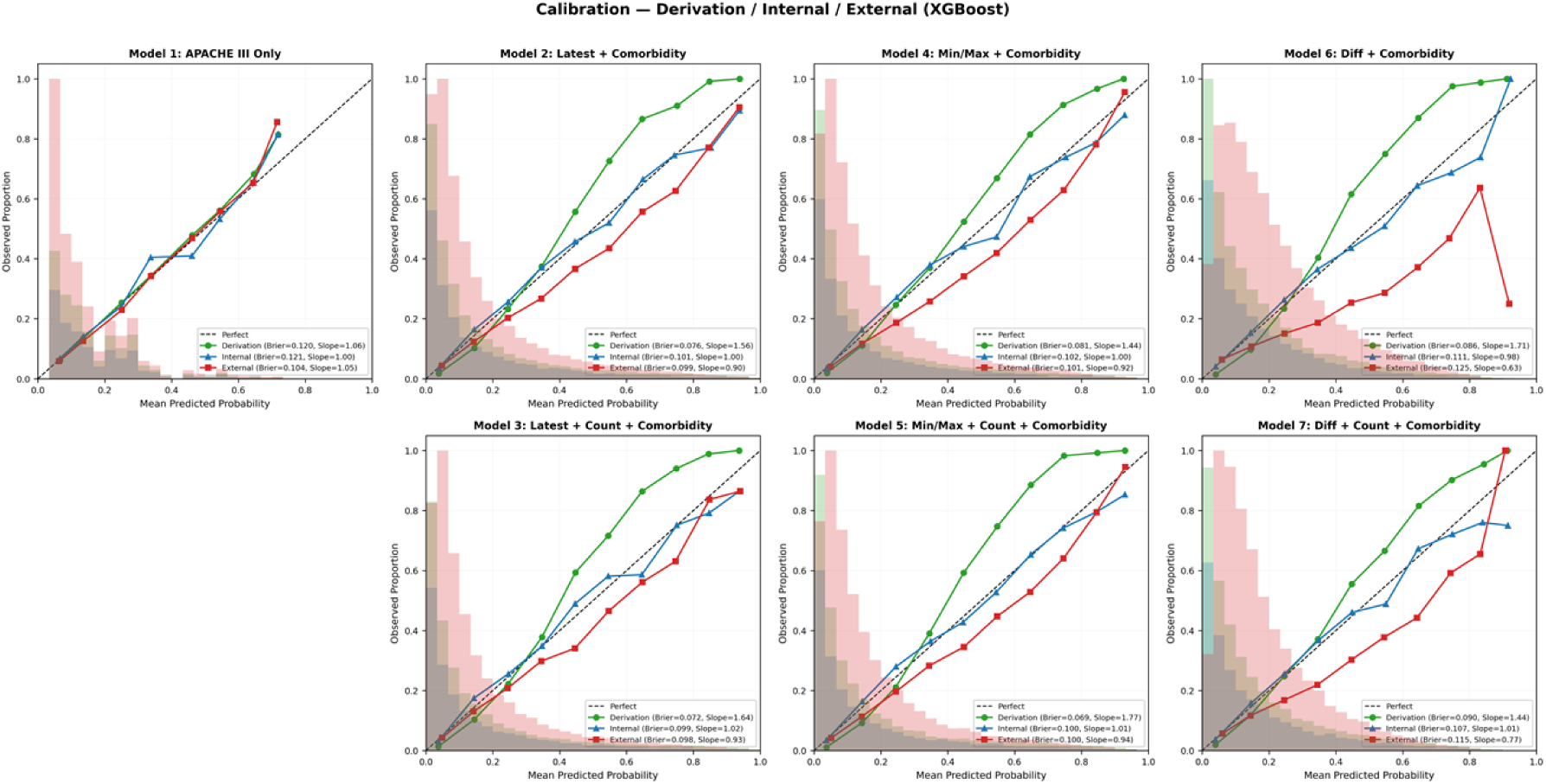
Calibration plots for XGBoost models on derivation, internal validation, and external validation sets Calibration plots for XGBoost Models 1–7 on derivation (training), internal validation, and external validation sets. XGBoost models maintain better calibration on external validation compared to logistic regression, particularly for models with count features.

In XGBoost, external calibration also tended to worsen with increasing complexity. For example, Model 1 had slope 1.046 and intercept −0.003, whereas Model 6 had slope 0.634 and intercept −0.984. In contrast to logistic regression, differences between paired specifications with and without measurement count features were small in the latest-value and min/max models: Model 2 versus Model 3 (slope 0.898 vs 0.927; intercept −0.373 vs −0.307) and Model 4 versus Model 5 (slope 0.918 vs 0.940; intercept −0.436 vs −0.375) (Table 3).

#### Subgroup analysis

We evaluated discrimination by race/ethnicity for Models 1 to 3 in the internal validation (MIMIC-IV) and external validation (eICU-CRD) cohorts (S7 Fig). In MIMIC-IV, subgroup sizes and event counts were Black n = 1,040 (153 deaths, 14.7%), Other n = 2,949 (637 deaths, 21.6%), and White n = 8,099 (1,181 deaths, 14.6%); in eICU-CRD they were Black n = 2,988 (438 deaths, 14.7%), Other n = 2,786 (391 deaths, 14.0%), and White n = 25,483 (3,520 deaths, 13.8%). From internal validation to external validation, the AUC decline was largest in the White subgroup for Models 2 and 3, whereas Model 1 showed similar AUC values between cohorts across subgroups (S7 Fig).

## Discussion

### Summary of key findings

In this study, we developed and externally validated prediction models for in-hospital mortality in ICU patients with sepsis, using a controlled set of prespecified model specifications to quantify how feature engineering choices are associated with domain shift. By holding the outcome definition, study cohorts, and modeling framework constant and varying only the representation of routinely collected physiologic variables and the inclusion of observation-process features, we evaluated the incremental association of specific feature choices with generalization performance. Internally, discrimination was higher for specifications using more detailed physiologic summaries (most recent values, extreme values, and within-window variability) and increased further when measurement counts were added. In external validation, however, performance drops were larger for specifications using more complex physiologic representations. Models incorporating measurement counts exhibited larger declines in discrimination and worse calibration than their counterparts without counts, with the clearest degradation observed for logistic regression. XGBoost showed smaller incremental degradation from measurement counts in the latest-value and min/max specifications, although complex specifications still exhibited marked domain shift. These findings indicate that feature engineering decisions, particularly the inclusion of observation-process variables, are associated with a tradeoff between internal performance gains and external transportability.

### Comparison with prior research

Our study design, which held the outcome and evaluation framework constant across two independent databases, allowed us to show that transportability can deteriorate even when internal performance improves, identifying feature engineering as a key and modifiable factor associated with cross-setting degradation. This finding aligns with prior work demonstrating that routinely collected clinical data reflect not only underlying patient state but also how, when, and why measurements are obtained, and that these observation patterns can be locally informative yet unstable across settings [9,10,13]. Extending this line of research, we provided empirical evidence that increasing feature complexity, particularly through the addition of observation-process and within-window summary features, coincides with larger external performance drops and more pronounced miscalibration.

Our findings are also consistent with evidence that observation-process features, including ordering patterns and measurement frequency, can carry strong predictive signal, sometimes rivaling the predictive value of the measured results themselves [14,16]. Meanwhile, external validation studies have highlighted that apparently strong development performance can mask unreliable risk estimation after deployment, as illustrated by the independent evaluation of the Epic Sepsis Model [6]. In this context, our results add deployment-relevant evidence: feature sets enriched with observation-process signals may be associated with improved internal discrimination alongside larger cross-dataset degradation, with calibration providing a particularly sensitive indicator of reduced transportability.

### Mechanisms underlying external calibration degradation

The observed calibration degradation can be explained by the fact that extreme-value summaries, within-window variability, and measurement intensity partially encode setting-specific observation and care processes in addition to physiology. Measurement intensity is shaped by staffing ratios, monitoring protocols, documentation practices, and institutional culture, so its distribution can differ substantially across sites even among patients with similar clinical severity [9,14]. When models learn relationships between these features and outcomes in the derivation setting, they can produce systematically miscalibrated predictions when applied elsewhere, consistent with our observation that specifications including measurement counts showed lower calibration slopes and more negative calibration intercepts on external validation. Notably, some distributional differences between databases may also reflect variable harmonization challenges rather than observation-process differences alone; for example, the large difference in mean FiO2 between MIMIC-IV (92.5%) and eICU-CRD (59.4%) likely reflects recording conventions rather than true clinical differences, and may contribute to domain shift in all specifications that include FiO2-derived features. However, because FiO2 was included in all specifications (Models 2–7), this variable affects all models equally and does not explain the differential degradation pattern across specifications. This interpretation is further supported by the marked distributional differences in measurement counts between the two databases (S2 Fig) and by the finding that variability-based specifications (max–min) exhibited the largest cross-dataset degradation, suggesting that within-window range summaries capture monitoring patterns that are less stable across institutions than the underlying physiologic values from which they are derived. Importantly, a zero measurement count does not distinguish between variables that were not clinically indicated and variables that were indicated but not recorded. Among the variables in our study, vital signs such as heart rate and respiratory rate are universally monitored in ICU patients, and a zero count almost certainly reflects a documentation gap; in contrast, arterial blood gases, bilirubin, and albumin are ordered selectively based on clinical indication, and a zero count may reflect an appropriate clinical decision. If the relative frequency of these mechanisms differs across sites, the zero-count encoding could itself drive domain shift independently of the observation-process mechanism we describe [26–28].

An alternative explanation is that the observed degradation reflects overfitting due to increased dimensionality rather than the specific nature of the added features. However, dimensionality-matched comparisons partially address this concern: Model 2 and Model 6 had the same number of input features (21 each) yet showed markedly different external degradation (ΔAUROC −0.047 vs −0.098 in logistic regression), suggesting that the type of physiologic summary, not merely the number of inputs, is associated with domain shift magnitude.

A related alternative explanation is that the observed degradation reflects overfitting in the more complex specifications rather than sensitivity to cross-setting differences. However, the pattern of external miscalibration was not aligned with internal optimism alone. Logistic regression showed only small training-to-internal-validation performance gaps yet exhibited the most pronounced external calibration deterioration when measurement count features were included. In contrast, XGBoost showed larger internal optimism but smaller incremental degradation from count features in several paired comparisons. This dissociation between internal overfitting metrics and external calibration failure, together with the marked distributional shifts in measurement counts between databases (S2 Fig), is more consistent with differences in the data-generating process for observation-process features than with overfitting alone.

### XGBoost robustness relative to logistic regression

The smaller incremental degradation from measurement count features observed in XGBoost may reflect several mechanisms. Gradient-boosted trees use binary splits that effectively threshold count features into discrete ranges; if the clinically relevant distinction is between zero and non-zero measurements, these thresholds may remain valid even when exact count distributions shift across sites. In contrast, logistic regression assigns a single linear coefficient to each count feature, so any distributional shift propagates directly through the predicted log-odds [25]. Additionally, the ensemble structure and regularization of XGBoost (maximum depth constraints, subsampling, and L1/L2 penalties) can limit the influence of any individual shifted feature. These mechanisms are consistent with evidence that covariate shift corrections can improve generalizability of sepsis prediction models [29], and with our observation that XGBoost showed minimal incremental degradation from count features in the latest-value and min/max specifications while still exhibiting substantial domain shift in variability specifications where the underlying physiologic features themselves were less stable. Future work incorporating feature importance analyses could empirically verify which mechanism predominates.

### Subgroup differences in transportability

The larger AUROC decline observed in the White subgroup may reflect several factors. Because White patients comprised the largest subgroup in both cohorts, cross-setting degradation was more precisely estimated, with less sampling variability potentially masking the shift in smaller subgroups. In addition, observation-process shift may have been more strongly expressed in this group if measurement-intensity patterns differed more substantially between MIMIC-IV and eICU-CRD for White patients than for other racial subgroups. These subgroup differences are descriptive and hypothesis-generating; they warrant targeted evaluation in future multi-site studies with adequate subgroup sample sizes to clarify whether transportability varies systematically by race and whether this variation is driven by differential observation-process encoding.

### Implications for model development and deployment

Our findings carry direct implications for the development and deployment of clinical prediction models. Observation-process features such as measurement counts were associated with improved internal discrimination but also with increased vulnerability to domain shift, potentially producing overconfident and miscalibrated predictions in new settings. This tradeoff suggests that feature selection for models intended for external use should include an explicit assessment of whether candidate features are likely to reflect stable pathophysiology or setting-specific observation and care processes. When observation-process features are included, their distributional stability across target deployment settings should be evaluated before implementation. Tree-based models appeared more resilient to this type of domain shift than logistic regression in our analysis, but this relative robustness should not be assumed without empirical verification in each deployment context. More broadly, external validation should assess calibration alongside discrimination, as calibration degradation provided the earliest and clearest signal of reduced transportability across all model specifications.

### Limitations

This study has several limitations. First, the derivation cohort came from a single academic tertiary care center, so fitted relationships and feature behavior may reflect local case mix, clinical workflows, and documentation practices, even though external validation was performed in a large multi-center dataset spanning 208 hospitals. Second, despite efforts to harmonize variables across databases, differences in recording conventions may have introduced non-equivalent variable definitions (e.g., FiO2 values). Because such harmonization artifacts affect all model specifications equally, they do not explain the differential degradation across specifications but may have contributed to a common baseline level of domain shift. Third, the derivation and validation periods only partially overlapped (2008 to 2022 versus 2014 to 2015), so temporal changes in clinical practice, sepsis management guidelines, and documentation standards could have affected predictor distributions and baseline risk independently of feature engineering choices. Fourth, we compared two model classes, logistic regression and XGBoost, and did not evaluate other approaches such as neural networks or domain adaptation methods that might mitigate observation-process shift. Fifth, we evaluated transportability in one direction only (single-center derivation to multi-center validation). This design reflects the common deployment scenario in which a model developed at one institution is applied elsewhere, and the 208-hospital eICU-CRD provided a stringent test of this scenario. Nevertheless, a reverse validation (training on eICU-CRD and validating on MIMIC-IV) would help disentangle observation-process feature instability from idiosyncratic characteristics of single-center derivation data, as demonstrated by bidirectional cross-database pipelines in critical care research [30]. Sixth, although dimensionality-matched comparisons suggest that feature type matters beyond dimensionality alone, we did not perform a formal null comparison to fully exclude the contribution of increased dimensionality. Finally, our analysis was restricted to adult ICU patients meeting Sepsis-3 criteria and in-hospital mortality as the outcome; the observed tradeoffs between internal performance and external transportability may not generalize directly to other populations, clinical settings, or prediction targets.

## Conclusion

Using a controlled experimental framework that evaluated the association between feature engineering choices and domain shift, we found that enriching prediction models with observation-process features and complex physiologic summaries was associated with improved internal discrimination but progressively worse external calibration and transportability. Measurement count features, proxies for measurement intensity, were consistently associated with larger cross-dataset performance degradation, particularly in logistic regression. These findings underscore the need for model developers to critically evaluate whether candidate features encode stable pathophysiology or setting-specific care processes, and highlight the importance of calibration assessment in external validation as the most sensitive indicator of reduced transportability.

## Author contributions

**Conceptualization:** RY, FW, LKS, AA, LAC, TT. **Data curation:** RY, FW. **Formal analysis:** RY, FW. **Methodology:** RY, FW, LKS, AA, TT. **Software:** RY, FW. **Supervision:** TT. **Validation:** RY, FW. **Visualization:** RY. **Writing – original draft:** RY. **Writing – review & editing:** RY, FW, LKS, AA, LAC, TT.

## Supporting information

**S1 Fig. Predictor distributions.** Overlaid histograms comparing the distributions of each physiologic predictor variable between the derivation cohort (MIMIC-IV, blue) and the external validation cohort (eICU-CRD, orange). Each panel shows one of the 18 continuous physiologic variables plus age, APACHE III score, and urine output. Distributional differences are apparent for several variables, including FiO2, alveolar-arterial gradient, and urine output, reflecting cross-database differences in patient case mix, measurement conventions, and recording practices.

**S2 Fig. Measurement count distributions.** Overlaid histograms comparing the distributions of measurement counts (number of recorded measurements within the first 24 hours) for each physiologic variable between the derivation cohort (MIMIC-IV, blue) and the external validation cohort (eICU-CRD, orange). MIMIC-IV generally showed higher measurement frequencies across most variables. These distributional differences in observation-process features are a key hypothesized driver of the domain shift observed when measurement count features were included in prediction models.

**S3 Fig. ROC curves for logistic regression.** Receiver operating characteristic curves comparing internal validation (blue, solid) and external validation (orange, dashed) performance for logistic regression Models 1–7. Upper row: models without measurement count features (Models 1, 2, 4, 6); lower row: paired models with measurement count features (Models 3, 5, 7). AUC values are shown in each panel. The gap between internal and external curves widened with increasing feature complexity and with the addition of measurement count features.

**S4 Fig. Precision-recall curves for logistic regression.** Precision-recall curves comparing internal validation (blue, solid) and external validation (orange, dashed) performance for logistic regression Models 1–7. Average precision values are shown in each panel. Upper row: models without measurement count features; lower row: paired models with measurement count features. External average precision declined progressively with more complex specifications, with larger drops observed for models incorporating measurement counts.

**S5 Fig. ROC curves for XGBoost.** Receiver operating characteristic curves comparing internal validation (blue, solid) and external validation (orange, dashed) performance for XGBoost Models 1–7. Upper row: models without measurement count features; lower row: paired models with measurement count features. AUC values are shown in each panel. XGBoost showed smaller incremental degradation from measurement count features compared with logistic regression in the latest-value and min/max specifications, although complex specifications (Models 6 and 7) still exhibited substantial domain shift.

**S6 Fig. Precision-recall curves for XGBoost.** Precision-recall curves comparing internal validation (blue, solid) and external validation (orange, dashed) performance for XGBoost Models 1–7. Average precision values are shown in each panel. Upper row: models without measurement count features; lower row: paired models with measurement count features. External average precision declined with increasing specification complexity, consistent with the AUROC-based findings.

**S7 Fig. Subgroup performance by race/ethnicity.** Receiver operating characteristic curves stratified by race/ethnicity (White, Black, Other) for Models 1, 2, and 3, comparing internal validation (MIMIC-IV, blue, solid) and external validation (eICU-CRD, orange, dashed) performance. Each panel shows AUC and sample size for one model–subgroup combination. Model 1 (APACHE III only) showed stable AUC across cohorts in all subgroups. Models 2 and 3 showed larger AUC declines in external validation, with the most pronounced drop observed in the White subgroup.

**S1 Table. Model feature summary.** Complete list of predictor variables for each model specification.

**S2 Table. Recalibration-only results.** External validation recalibration metrics across model specifications and algorithms.

## Notes

### Competing Interest Statement

The authors have declared no competing interest.

